# Mathematical modeling of the waning of anti-RBD IgG SARS-CoV-2 antibody titers after a two-dose BNT162b2 mRNA vaccination

**DOI:** 10.1101/2022.08.15.22278583

**Authors:** Javier Torres, Jesús Ontañón, Carlos de Cabo, Julia Lozano, María Ángeles Requena, Joaquín Blas, José Luis Rodríguez-García, Antonio Mas, Francisco J. Cimas, Javier Solera

## Abstract

**Objective:** Serum antibody levels have been linked to immune protection in SARS-CoV-2 infection. After exposure to the virus and/or vaccination there is an increase in serum antibody titers followed by progressive non-linear waning of antibody levels. Our aim was to find out if this waning of antibody titers would adjust to a mathematical model.

**Methods:** We studied serum anti-RBD (receptor binding domain) IgG antibody titers over a ten-month period in a cohort of 54 health-care workers who were either never infected with SARS-CoV-2 (naive, nHCWs group, n = 27) or previously infected (experienced, eHCWs group, n = 27) with the virus after the second dose of the BNT162b2 mRNA vaccine. We have selected a risk threshold of 1000 UA/ml anti-RBD Ab titer for symptomatic infection based on the upper titer threshold for those volunteers who suffered infection prior to the omicron outbreak. Two mathematical models, exponential and potential, were used to quantify antibody waning kinetics and the relative quality of the goodness of fit to the data between both models was compared using the Akaike Information Criterion.

**Results:** We found that the waning of anti-RBD IgG antibody levels adjusted significantly to both exponential and potential models in all participants from both the naïve and experienced groups. Moreover, the waning slopes were significantly more pronounced for the naive when compared to the experienced health-care workers. In the nHCWs group, titers would descend below this 1000-units threshold at a median of 210.6 days (IQ range: 74,2). However, for the eHCWs group, the risk threshold would be reached at 440.0 days (IQ range: 135,2) post-vaccination.

**Conclusions:** The goodness of the fit of the anti-RBD IgG antibody waning would allow us to predict when the antibody titers would fall below an established threshold in both naive and previously infected subjects.

## Introduction

COVID-19 pandemic has represented a huge challenge for societies and health systems all over the world. While there are more and more people with an immune shield, either because they had recovered from primary SARS-CoV-2 infection, they have been vaccinated or both (1), the virus has continued to evolve with the emergence of new genetically distinct variants that imply a higher transmission rate and a decrease in the immune protection against re-infection (2-4)

Vaccines against SARS-CoV-2 have demonstrated a high degree of protection against Covid19 over time, at least in terms of severe disease and mortality (5,6). However, immunity to SARS-CoV-2 declines in a nonlinear fashion (7,8) and makes it difficult to estimate risk and take decisions about when to schedule booster vaccines (9). In fact, it has already been described that antibody (Ab) titers correlate with immune protection, and it has been used to predict protection against infection (10-14). This decrease in protection is different in vaccinated subjects without previous infection (naïve) and previously-infected (experienced) vaccinated subjects due to hybrid immunity (1,15-16). In this work, we analyzed the anti-RBD IgG Ab titer and the breakthrough infections from February 2021 to December 2021, just before the third vaccination dose, in a cohort of HealthCare Workers (HCW) vaccinated with RNA BNT162b2 (Pfizer Biotech.) vaccine in January 2021. We propose that it is possible to model the waning in the level of anti-RBD Ab titer over time through simple mathematical models. We have used the exponential and potential models, which are accurate models to analyze the non-linear waning of antibody titers for both natural infection and vaccination (11, 17-20). Our aim is to develop a simple way to describe the evolution of the antibody titer over time. This would allow us to predict a personalized optimal moment of administration of the booster vaccine dose as well as estimate the risk of infection and its severity according to an established threshold (correlate of protection).

## Methods

### Study design and oversight

We conducted an observational prospective longitudinal study continuing from a previously reported HCWs cohort from the Albacete General Hospital (CHUA, Spain) (21). This study was officially approved by the *Comité de Ética de la Investigación con medicamentos (CEIm) de la Gerencia de Atención Integrada de Albacete* (Internal code: 2021-12 EOm). Written informed consent was obtained from all study participants. These HCWs were vaccinated in January 2021 with the BNT162b2 mRNA vaccine. In brief, 63 HCWs from the original CHUA cohort volunteered to measure their antibody levels at several time-points during three periods. First, before the onset of vaccination; second, 7, 14 and 21 days after each of the two doses and third, monthly up to the administration of the third dose of the vaccine (December 2021). Of the 63 original HCW, we selected 54 subjects that complied with the follow-up schedule; at least 4 decreasing consecutive measurements after completing the vaccination schedule. Subjects were classified into two groups: naïve health-care workers (nHCW), which included participants without clinical or laboratory data suggestive of infection with the SARS-CoV-2 virus prior to vaccination, and experienced health-care workers (eHCW), consisting of those with previous SARS-CoV-2 infection. The kinetics of antibody decay was evaluated individually. For each subject, the point of maximum antibody level (between 7 and 45 days after second dose vaccination) and all those corresponding to the determinations made during the following 10 months were taken. We monitored the eventual appearance of breakthrough symptomatic or asymptomatic infections. Breakthrough infections were defined as the detection of SARS-CoV-2 by PCR 14 or more days after receiving the second dose.

### Biochemical analysis

Total IgG antibody levels against the S1 subunit of the SARS-CoV-2 virus spike protein that binds to the receptor binding domain (RBD) were measured using the SARS-CoV-2 IgG II Quant immunoassay in the ARCHITECT i-System (Abbott, Abbott Park, US). The analytical measurement range is from 21 to 80,000 AU/mL and we used the manufacturer’s recommended cutoff point of 50 AU/mL for determining positivity. To convert AU/ml into international standard WHO units the conversion factor is 1/7 (22). To assess re-infection detection, PCR was performed in samples of nasopharyngeal exudates that were collected in tubes with 3 ml of universal transport medium (UTM) without inactivation and routinely sent to our laboratory for diagnosis of SARS-CoV-2. Samples were extracted with MagMaxTM Viral/Pathogen Nucleic Acid Isolation Kit (ThermoFisher) reagents using the KingFisher extractor (ThermoFisher) following manufacturer instructions. For the qualitative detection of SARS-CoV-2 nucleic acid, the commercial TaqPathTM COVID-19 CE-IVD Kit was used together with the ThermoFisher QuantStudio 5 (QS5) thermal cycler.

### Statistical analysis

Quantitative demographic variables were expressed as mean and range, mean and standard deviation (SD) and confidence interval 95% (CI). Qualitative variables were expressed as number and percentage. Total anti-SARS-CoV-2 spike RBD region IgG antibody levels were reported using geometric mean concentrations (GMC).

Normality of the distributions were tested using Lilliefors test and variances between populations using the F-test. The two-tailed U-Mann-Whitney non-parametric method was used to compare different means between the nHCW and eHCW groups. Within-group differences in total IgG levels obtained at the different time points were assessed using the two-tailed Wilcoxon Sign test. Chi-square and Fisher’s test were used to compare categorical data. All the confidence intervals, as well as the statistical tests, were calculated with a significance level of 95%.

Two mathematical models were used to quantify antibody waning kinetics. First, the exponential model *y = A·e^^^(t·x)* was used for each patient, where *y* is the SARS-CoV-2 RBD IgG antibody concentration, *A* is the exponential transformation of the extrapolated SARS-CoV-2 IgG concentration of the HCW at day 0, *t* is the slope of the model and *x* is the time after vaccination in days. The parameters of the model were estimated by fitting a linear model to the Naeperian logarithm of the SARS-CoV-2 RBD IgG concentration versus the time after vaccination expressed in days, using the Ordinary Least Squares method. The exponential curve is afterwards obtained by reversing the logarithmic transformation.

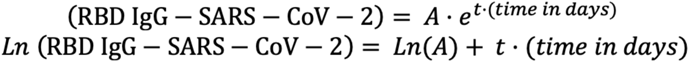

### Equation 1: Exponential and linear representations of the concentration of IgG versus time

A potential model was also used, represented by the curve *y = A · x^^^t* employing the same variable definitions as before. The adjustment was performed by Ordinary Least Squares regression of the Napierian logarithm of the concentration of SARS-CoV-2 IgG versus the Napierian logarithm of time. Exponential transformation was performed to obtain the curve from the linear adjustment:

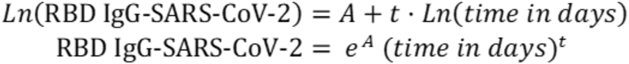

### Equation 2: Potential and linear representations of the concentration of IgG versus time

Relative quality of the goodness of fit to the data between both models was compared using the Akaike Information Criterion (AIC) (23). AIC was calculated for each patient and model. The difference of the AIC value was calculated as the exponential model’s AIC minus the potential model’s AIC. Positive delta values were interpreted as a better exponential fit and negative values as a better potential fit.

An ideal waning curve was built for each mathematical model and sub-cohort using the corresponding mean values for each group.

The calculations were carried out with the statistical software R, version 4.0.2 and data visualization figures were drawn with the ggplot2 package.

## Results

### 1- Characteristics of the Study Population

54 HCWs were included in the study, of which 27 were nHCW and 27 eHCW regarding previous infections with SARS-CoV-2 before vaccination. The epidemiological characteristics of the participants are described in Table 1. No significative differences were found regarding age, sex, number of Ab-titer determinations (timepoints) and follow-up duration between the two groups.

**Table 1.** Demographic characteristics of the participants in the study: *U-Mann-Whitney test;

|  | nHCW | eHCW | p-value |
| --- | --- | --- | --- |
| **Number** | 27 | 27 |  |
| **Age in years. Mean ± SD (Range)** | 42.04±14.91 (25-62) | 47.96±12.78 (25-67) | 0,12 * |
| **Number of females (%)** | 19 (70%) | 19 (70%) | 1.00 ** |
| **Follow-up days after the second dose of the vaccine. Mean ± SD (Range)** | 293.52 ± 3.50 (21-27) | 293.07 ± 10.82 (21-28) | 0.84 * |
| **Number of determinations per subject. Mean ± SD (Range)** | 6.85 ± 2.35 (4-12) | 6.33 ± 1.33 (4-10) | 0.32 * |

### 2- A differential decrease in Ab titers after the vaccine was observed between nHCWs and eHCWs

The geometric mean of the maximal post-vaccine values for the eHCW group was two times higher than the nHCW group (46.682 AU/ml Vs 23.623 AU/ml, p-value < 0.001).

We plotted the individual Ab waning curves from the measured Ab titers for each HCW using both exponential and potential models (Supp. 1). We observed a decrease in the anti-RBD Ab titers from the maximum post-vaccine values over the following 10 months in both the naïve and eHCW groups (Fig. 1A and 1B). The slopes of the curves for the nHCWs were more pronounced when compared with the eHCWs in both the exponential and potential models.

**Fig. 1.**
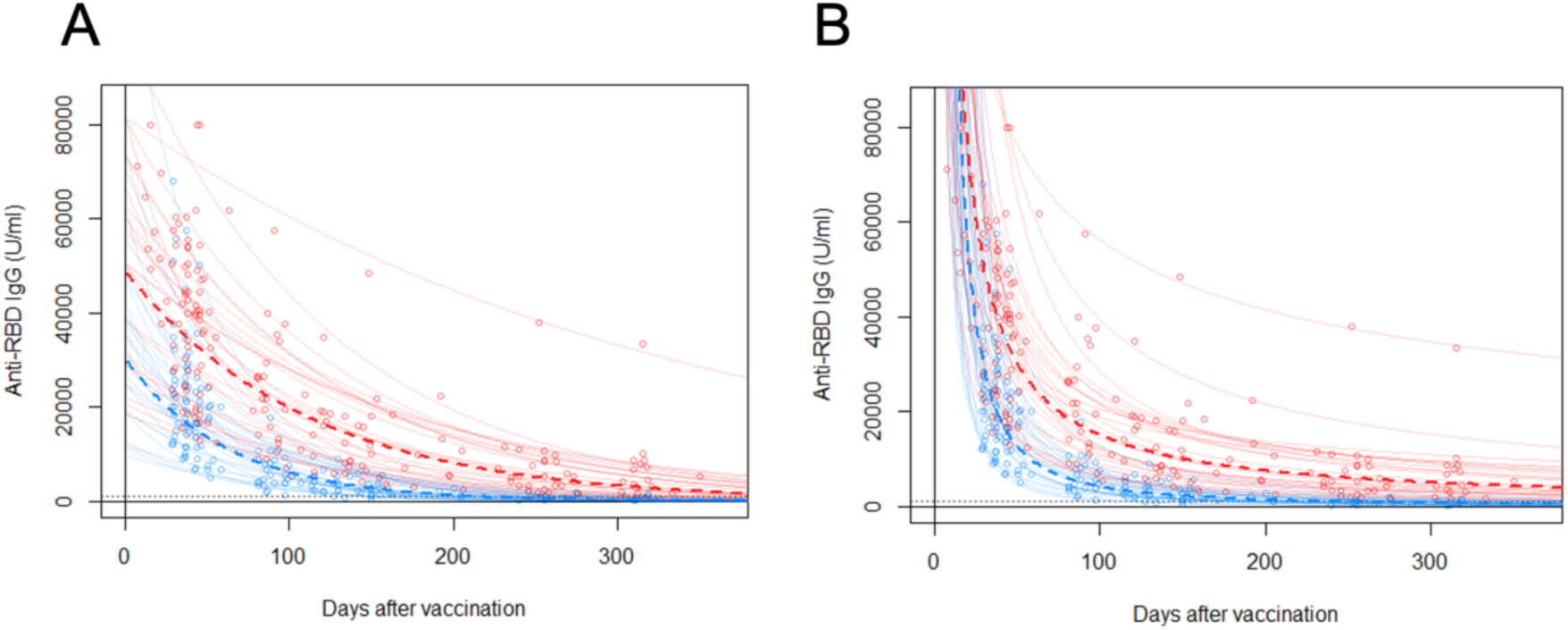
Time-course of anti-RBD IgG antibody titers for each individual HCW and the mean curves for exponential (A) and potential (B) models. Individual timepoints for each HCW are represented as open circles (red, eHCWs and blue, nHCWs). Each continuous line represents an individual antibody waning ideal curve for individual HCWs; dashed lines represent the ideal mean curves. The horizontal dashed line indicates 1.000 AU/ml.

First, for each HCW the individual data were fitted to an exponential curve, applying a linear regression method. Within the nHCWs group, the decay rate *t* had a median value of −0.015736 (Interquartile range: −0.019122 to −0.012970) and a mean value of −0.016137 (SD: 0.00412; 95% CI: −0.017692 to −0.014582). The R^2^ values for all the adjustments were between 0.899 and 0.996, with an average of 0.957 which demonstrates a good fit of the curve for every individual to their experimental data. All the p-values for the adjustments were significant (p-value range: 9.18E-7 to 5.0E-3) (Fig. 1A).

For the eHCWs group, the median *t* parameter was −0.008496 (Interquartile range: −0.009983 to −0.007256) and the mean value was −0,009054 (SD: 0,00299; 95% CI: −0.01018 to −0.0079). Their calculated curves had a good fit to the experimental data similar to the one found for the nHCW group (Fig. 1A). The R^2^ value for the adjustments was between 0.907 and 0.998, with an average value of 0.954. All the corresponding p-values were significant (p-value range: 1.406E-7 to 0.844E-3).

Second, in the case of the potential adjustment, for the nHCWs group, the parameter *t* corresponding to the linear regression slope when representing the *Ln* of the SARS-Cov-2 IgG concentration against the *Ln* of the time had a median value of −1.598 (Interquartile range: - 1.803 to −1.403) and a mean value of −1.623 (SD: 0.240; 95% CI: −1.713 to −1.523). A good fit of the curve to the experimental data was found for each individual (Fig. 1B). The R^2^ values for all the adjustments were between 0.970 and 0.999, with an average of 0.991. All the p-values for the adjustments were significant (p-value range: 1.31E-9 and 2.27E-3).

Regarding the eHCWs group, analysis showed a median parameter *t* of −0.977 (Interquartile range: from −1.161 to −0.877) and a mean value of −1.005 (SD: 0.247; 95% CI: −1.098 to −0.912). The calculated curves had a similarly good adjustment to the experimental data found for the nHCWs group (Fig. 1B). The R^2^ value for the adjustments was found to be between 0.931 and 0.9997, with an average value of 0.988. All the corresponding p-values were significant (p-value range: 4.91E-12 and 7.91E-3).

The slopes of the ideal curves for the nHCWs were more pronounced when compared with the eHCWs in the exponential (1.801E-9) and potential models (p-value: 9.399E-13), with a stronger significance in the last one (Fig. 2).

**Fig. 2.**
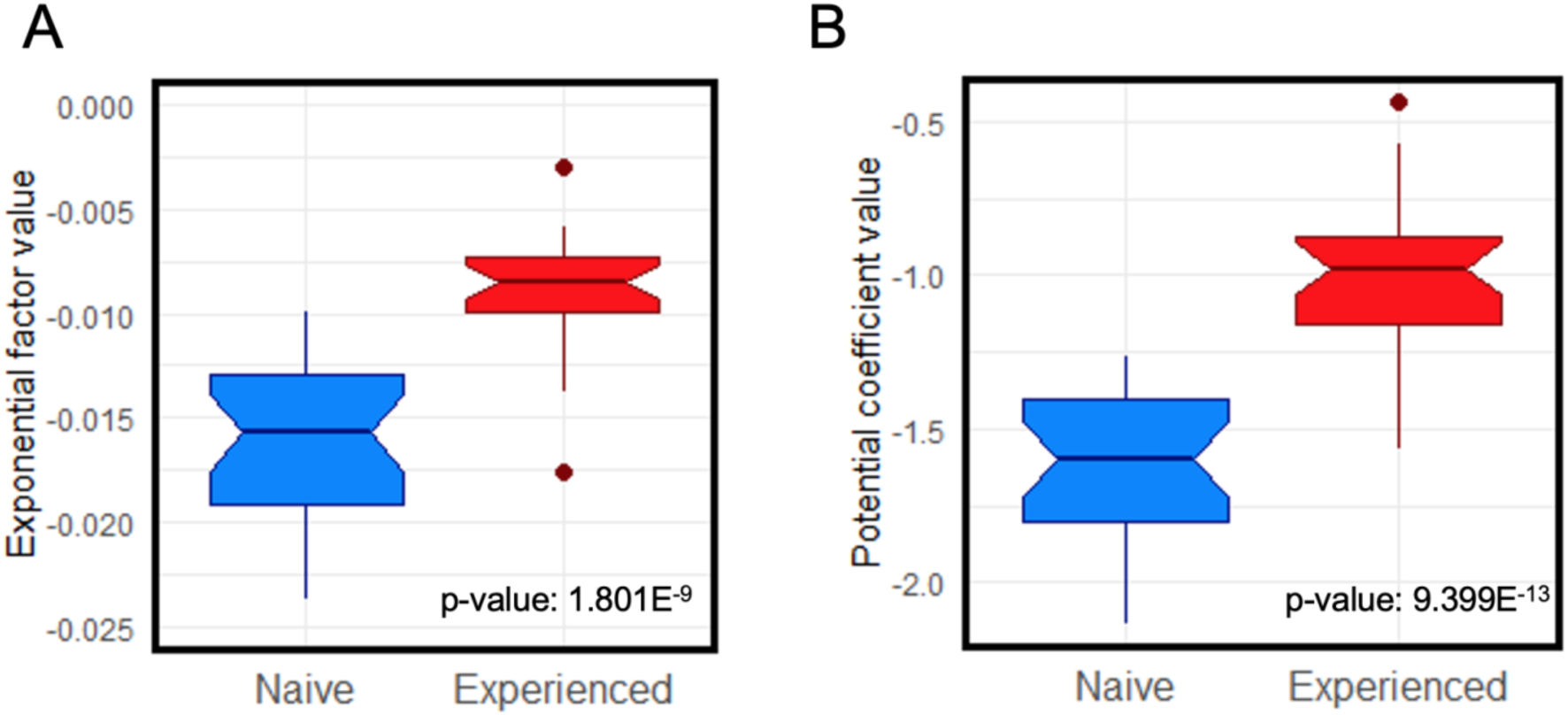
Notched box and whisker comparison of the mean of the ideal curves slope for exponential (A) and potential (B) models. Box width is proportional to the number of observations in each group (red, eHCWs and blue, nHCWs).

Comparison of both models was calculated for each individual HCW using the value of the AIC difference for the exponential minus potential model (deltaAIC) (Supp. 3). This difference was positive for 46/54 HCW (85.2%), denoting a better fit for the potential model. There were no statistically significant differences between the nHCWs and eHCWs according to the Fisher test (p-value: 0.704), denoting a better fit for the exponential model in both groups (Supp. Table 3).

### 3- Characterization of breakthrough infections in the studied cohort

Of the 54 HCWs included in this study, only 4 (7,4%) had breakthrough infections (Table 2). Although more of them (3) belonged to the nHCWs group and just 1 to the eHCWs group, no statistically significant differences were found (p-value = 0.6104). Two of nHCWs were infected in July 2021, when the predominant SARS-CoV-2 variant was delta and the other two infections happened in December 2021 when the predominant variant was Omicron. We estimated the anti-RBD antibody levels at the time of infection for all four HCWs. For the nHCWs the estimated value was < 1000 AU/ml, with a mean value of 504.2 AU/ml and a maximum of 932.6 AU/ml and a minimum of 15.0 AU/ml. For the eHCW, the anti-RBD antibody levels at the time of infection were < 4000 AU/ml (Table 2). The curve fitting R^2^ value for that subject was 0.955.

**Table 2.** Parameters of interest of the breakthrough infection cases detected during our study. Parameters include group, sex, existence of previous diagnosis, 2nd dose date, breakthrough infection date (in month/year format), anti-RBD IgG titer in breakthrough infection and adjusted R^2^ (calculated by exponential model/calculated by potential model).

| Subject | Group | Sex | Previous diagnosis | 2nd dose date | Break. inf. date | Titer on break inf. | $R^2$ |
| --- | --- | --- | --- | --- | --- | --- | --- |
| 54 | N | F | X | 02/2021 | 07/2021 | 563.9/1265.9 | 0.9715/0.9866 |
| 55 | N | M | X | 02/2021 | 07/2021 | 932.6/2276.6 | 0.9743/0.9764 |
| 48 | N | F | X | 02/2021 | 11/2021 | 16.0/527.8 | 0.9734/0.9956 |
| 26 | P | F | 03/2020 | 02/2021 | 12/2021 | 2960.5/4029.5 | 0.955/0.9769 |

### 4- Usefulness of the model to predict Abs levels and use of the data obtained in decision making

The post-vaccination half-life of the RBD-Ab titers estimated by the exponential model was 45.84 days on average (SD: 12.09, 95% CI: 41.28-50.41) for the nHCW group and, whereas for the eHCWs group, the half-life was 85.67 days (SD: 34.95; 95% CI: 72.48 to 98.85 days). This result for eHCWs is 1.87-fold higher than in the nHCWs group.

We established a preliminary risk threshold of 1000 UA/ml anti-RBD Ab titer for symptomatic infection based on the upper titer threshold for those volunteers who suffered infection prior to the omicron outbreak. In the nHCWs group, titers would descend this 1000-units threshold at 221.1 days post-vaccination, on average (SD: 53.1 days). The median for this group was 210.6 days (Interquartile range: 74,2). However, for the eHCWs group, the risk threshold would be reached at 483.3/38880 days post-vaccination, on average (SD: 229.1 days). For this group, the median is located at 440.0 days (Interquartile range: 135,2). The difference in the predicted mean time to descend to the risk threshold between nHCWs and eHCWs was statistically significant (p-value = 2.14E-12, Mann-Whitney U-test). Interestingly, in one individual from the eHCWs group a value below the 1000 UA/ml risk threshold would be reached as late as 1.468,9 days. Predictions made with the potential model develop an asymptotic behavior at long periods and are reflected on Supp. Table 2.

According to the extrapolations made, when 95% of the nHCWs group would have already fallen below this limit, only 5 (18.5 %) of the eHCWs would have fallen below this value, while the remaining 22 (81.5 %) would be still above. Similarly, when 50% of the eHCWs patients would be below the threshold, all the nHCW patients would already be below it.

## Discussion

In this study, we modeled the observed waning in anti-RBD IgG Ab levels in a cohort of HCWs following a second dose of BNT162b2 vaccine during 10 months by using two different mathematical approximations, the exponential and potential models. Both models adjust well, with strong correlation coefficients and statistical significance (Supp. Fig. 1). Thanks to the goodness of adjustment of the curves to either potential or exponential models, we would be able to predict individual Ab titers after the second those of the mRNA BNT162b2 vaccination over time by counting on just two Ab determination timepoints.

Interestingly, the potential model offers a better fitting for the early timepoints of determination, consistent with the quick waning in Ab titers during the first weeks after the peak levels. This observation is consistent with previous reports which describe a rapid decline in SARS-CoV-2 IgG Ab titer during the first four months after antigen contact, followed by a gentler waning over the following 7 months (11-12, 24). At this point, Ab levels correlate with the presence of antigen-specific plasma cells found in bone marrow (25). The potential model also presents a better adjustment than the exponential model according to the AIC criterion. A negative delta value was found for 85% of the subjects (Supp. Table 1). The exponential and potential models have been previously compared to describe anti-RBD IgG titer waning in a smaller, naïve cohort (11). That same study found a more robust fitting of the potential model for the anti-RBD Ab waning curve, similarly to our results. However, they found a better adjustment of the exponential model for other determinations of neutralizing Ab.

The nHCWs and eHCWs sub-cohorts have statistically significant different slopes, showing a pronounced decline for nHCWs in both models. Multiple studies have described that vaccinated subjects with previous infection have stronger and longer lasting response to vaccines than naïve ones, as well as lower risk of infection (1,6,15-16), which is attributed to hybrid immunity. Our results therefore agree with the proposed lower risk of infection for hybrid immunity bearers. Compared to previous works, our study provides a detailed Ab titer evolution throughout more timepoints and also demonstrates significant differences between the nHCWs and eHCW sub-cohorts slopes.

We found four subjects that had breakthrough infections detected during the follow-up of our cohort. All of them were mild and did not require hospitalization. Three of them corresponded to nHCWs and only 1 to eHCWs. The number of infections is higher for nHCWs as it has been widely described (1, 15-16). However, no statistical significance was found due to the reduced number of relapse cases given the small size of the cohort. With that in mind, those HCWs who experienced relapses had lower Ab levels at the time of infection. Conversely, the average Ab level of those who did not relapse was higher than those who did. The level of Abs against SARS-CoV-2 can be a marker of protection or risk (a correlate of protection) to determine individual risk of infection and optimal moment for vaccine booster. In this regard, testing our model in larger cohorts with more breakthrough infections will allow us to estimate more accurate risk of infection thresholds and correlates of protection.

Notwithstanding the above-mentioned goodness of fit of the models, we observed a high individual variability among the curves, that was more pronounced in nHCWs (Fig. 1) It is remarkable how, within the two well-differenced groups in terms of presence or absence of hybrid immunity, there is such a high heterogeneity in their anti-RBD Ab titer curves. This variability included subjects with a sustained low level in antibody titers (vulnerable to infection) while others were able to maintain high antibody titers over time, which would confer protection against infection. In particular, we found that one eHCW that presented such high sustained Ab titers that would not need a booster vaccination. These two types of populations of individuals according to their Ab response to vaccination have been well characterized by Nakamura et al. (24) in a larger cohort. The variability in the anti-RBD IgG antibody response supports the use of individual curves versus using a mean curve to attempt to predict Ab titers over time.

## Limitations

Our study was conducted in a small, homogeneous cohort of HCWs, who were mostly middle-aged and healthy subjects and therefore it may not represent the general population. Moreover, due to the period in which the study was performed, no new SARS-CoV-2 variants nor other vaccines or dose patterns were considered. Our model needs rigorous evaluation using data from different cohorts. Besides, antibody level estimations with our model for dates outside our period of study should be taken with caution. This model, like many other predictive models, is based on multiple regression techniques. In a recent study, this approach was combined with machine learning (19), which we did not use in our work. Nonetheless, in our study we do describe detailed individual waning curves comparing eHCWs with nHCWs at several timepoints and for a similarly long follow-up period (ten months).

## Conclusion

We described both exponential and potential models as parsimonious models that allow determination of the anti-RBD IgG Ab titer, building a personalized waning curve with only two antibody titer determinations. Therefore, we can estimate the moment when antibody titers drop below a certain threshold. Nevertheless, it should be noted that predictions would be most reliable within the observation period used in this work (10 months after the second dose of the vaccine). Regardless of the model used, eHCWs have a lower waning slope and longer persistence of antibody titers than nHCWs. Consequently, different vaccination booster schedules should be advised according to the individual persistence of antibody levels. Both the exponential and potential versatile models and allow the application of different thresholds that mark variables of clinical interest such as risk of hospitalization or symptomatic infection. Our modelization could also be used under different conditions which may alter conferred protection, such as new vaccines or viral variants. Our approach provides a tool for personalized predictions of Ab levels, and, thus, the rationalization of the administration of booster doses of anti-SARS-COV-2 vaccines, applying them only when necessary and avoiding potential side-effects.

## Data Availability

All data produced in the present study are available upon reasonable request to the authors

## Supplementary material

**Supp. 1.**
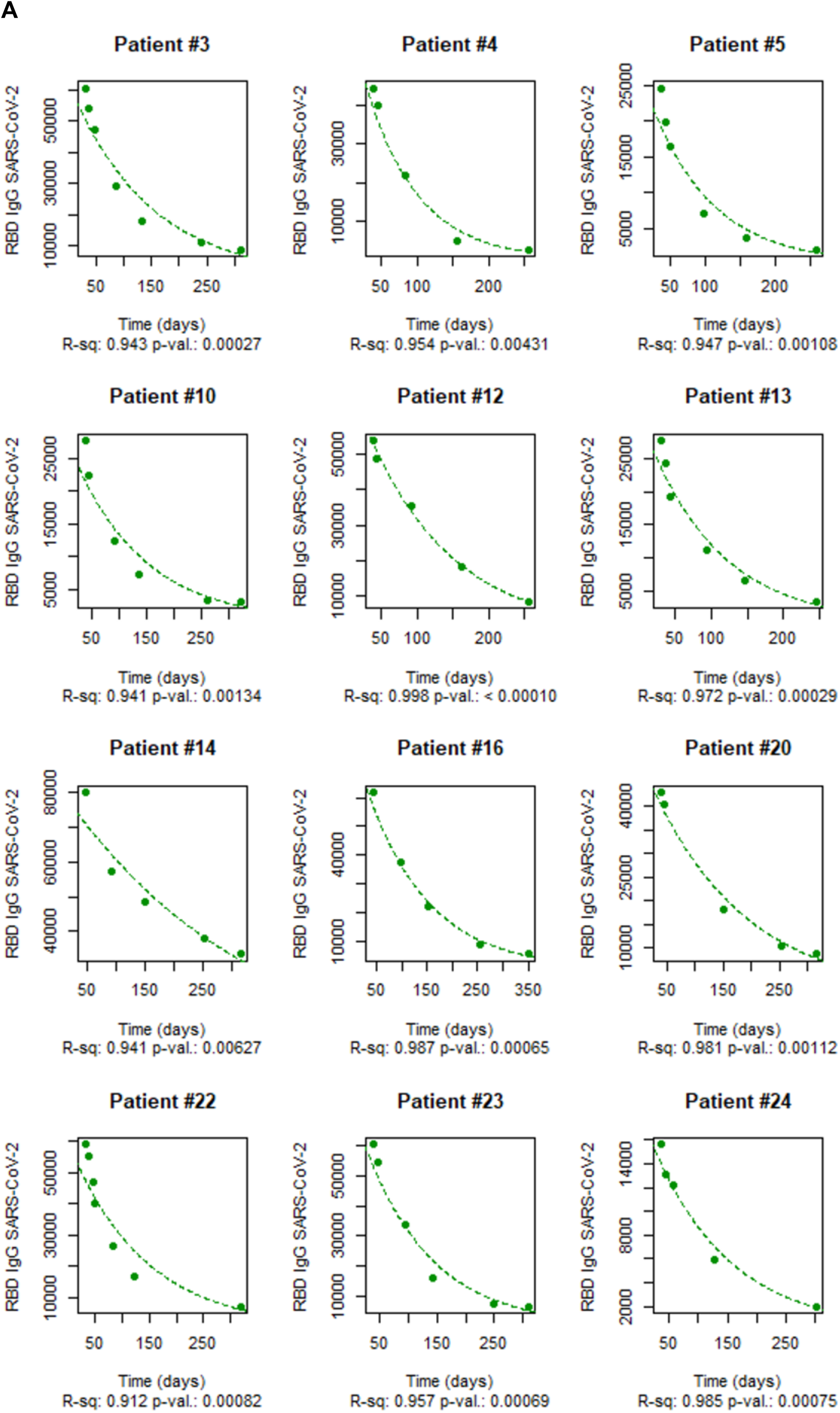
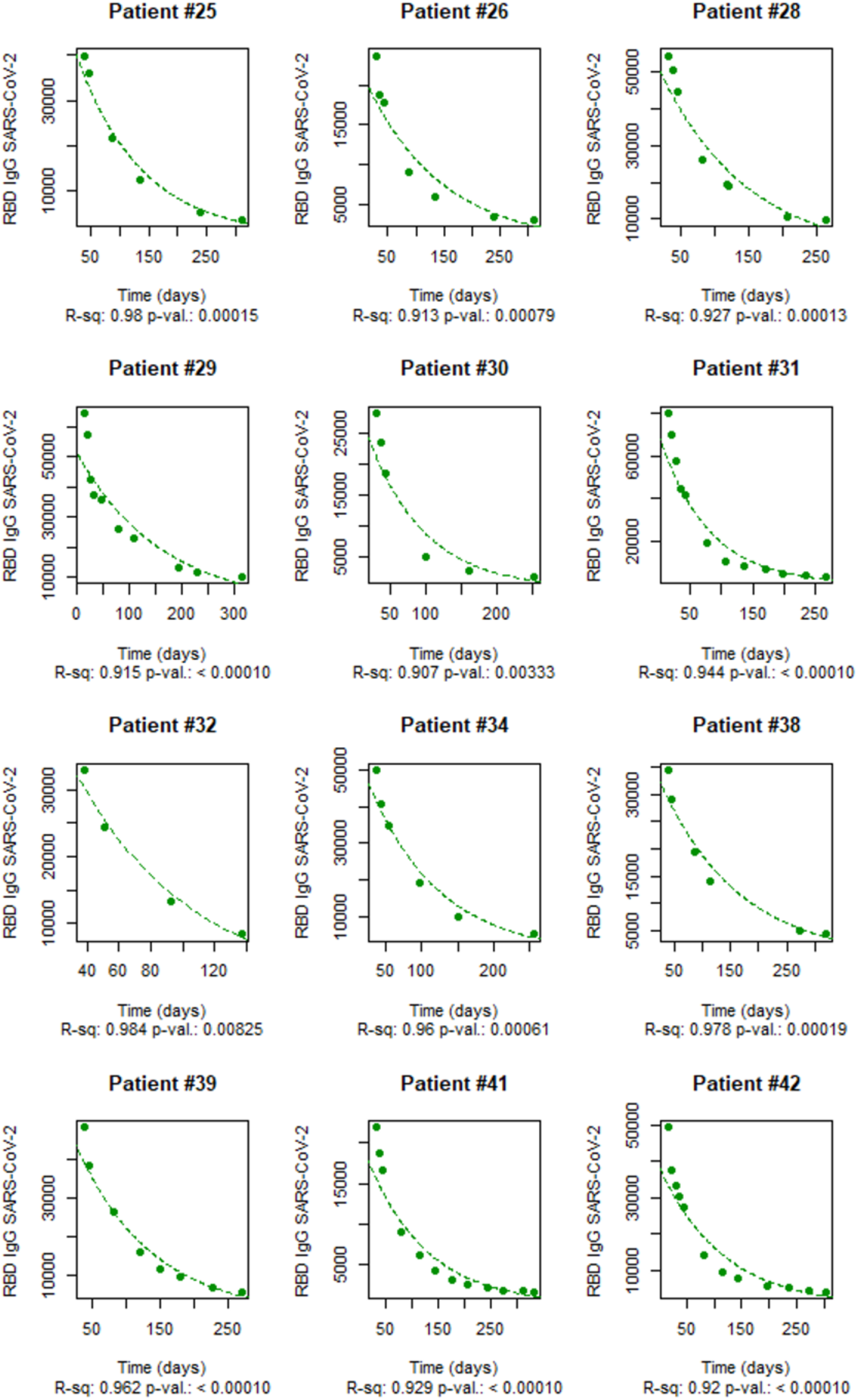
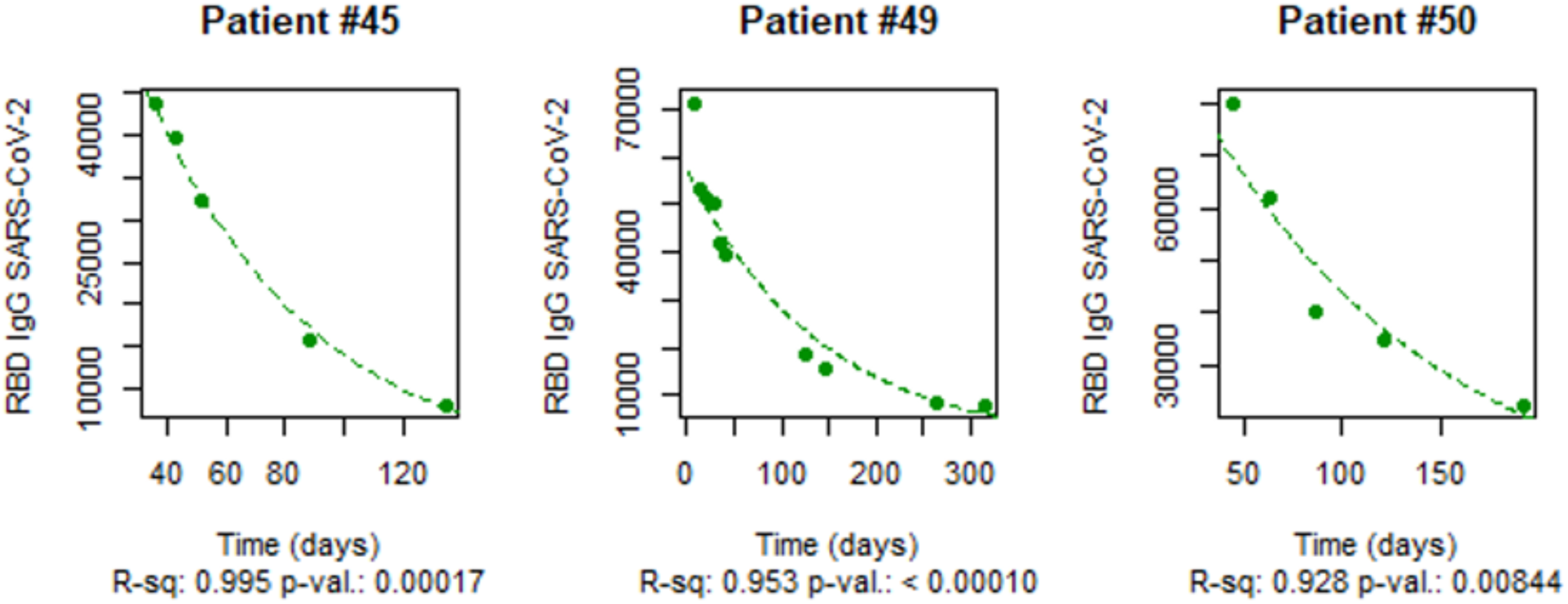
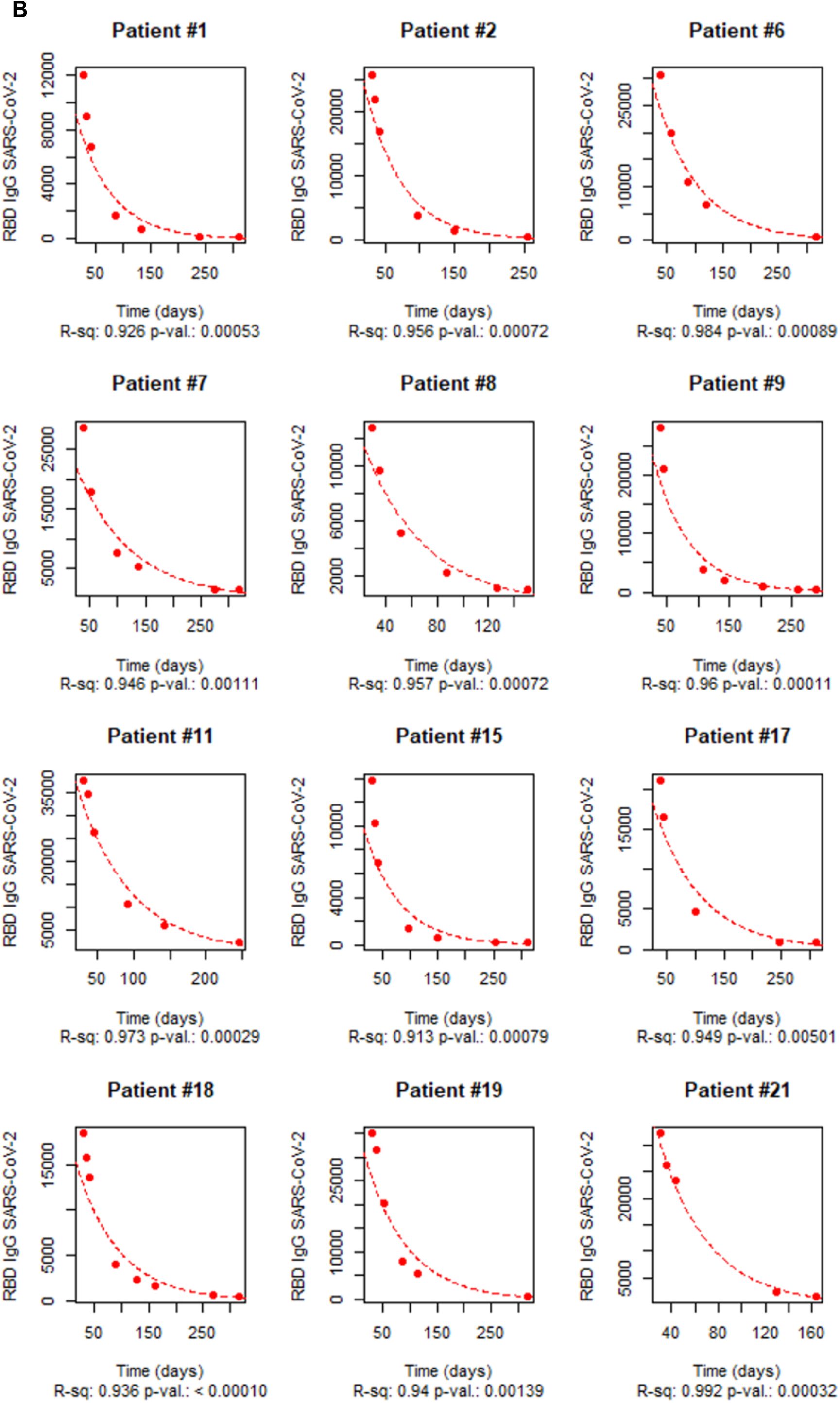
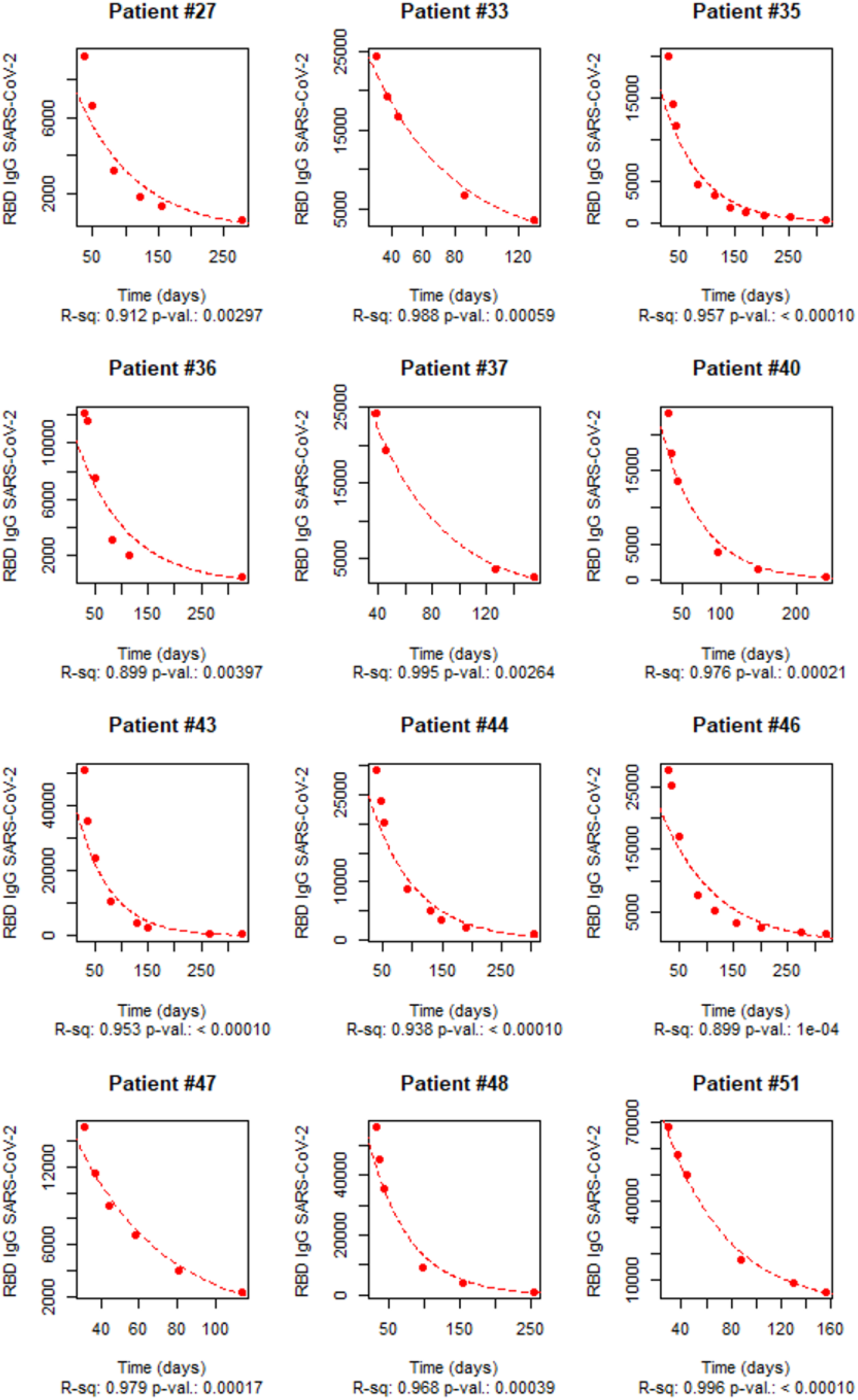
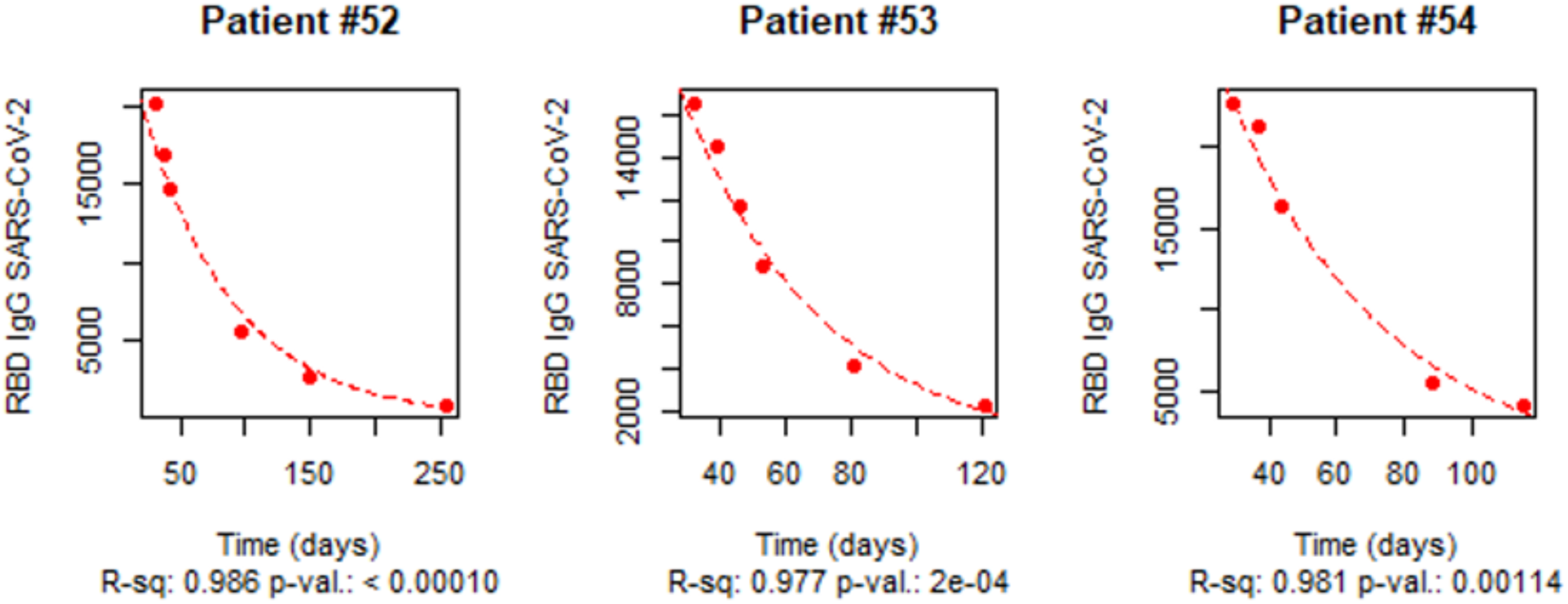
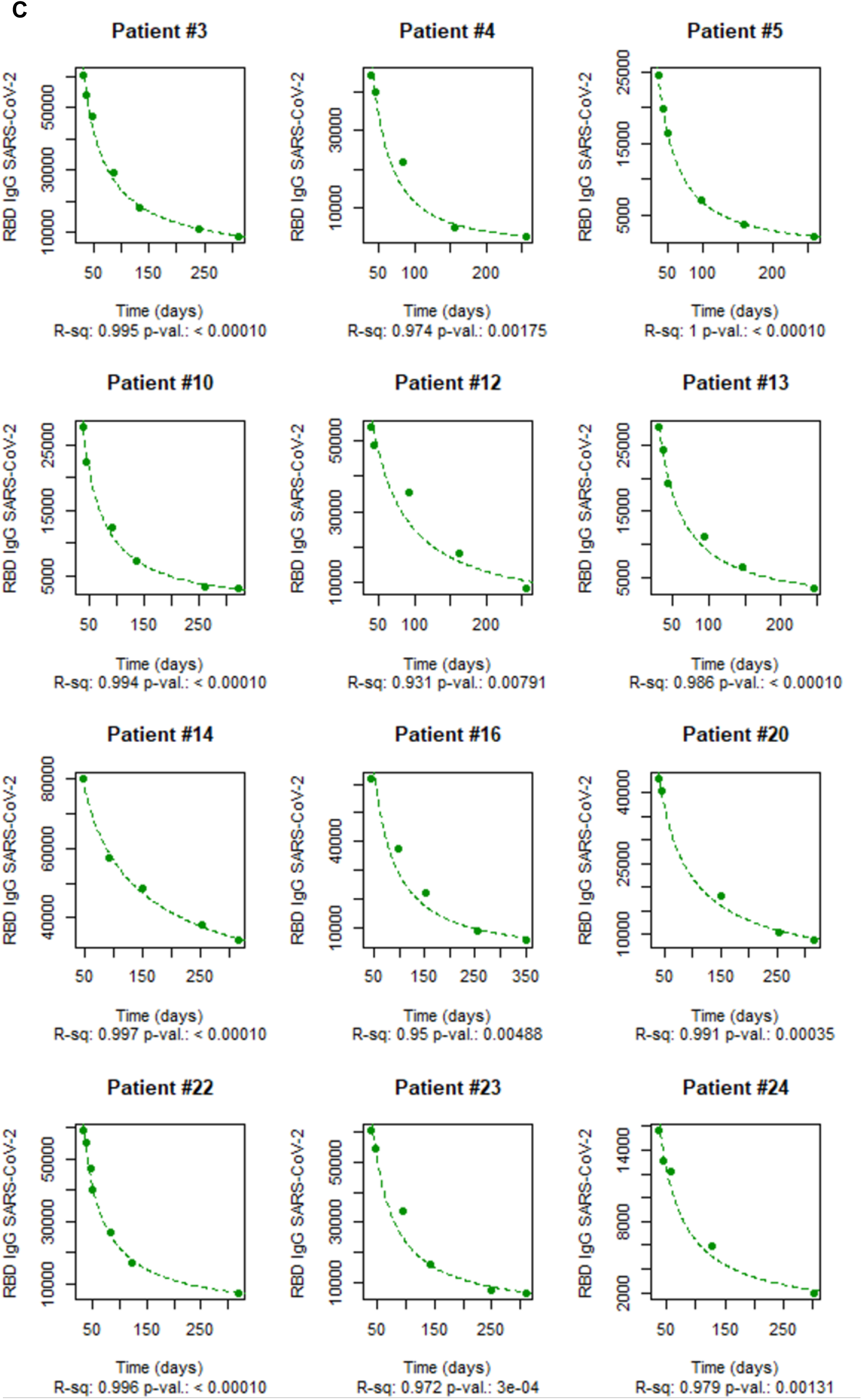
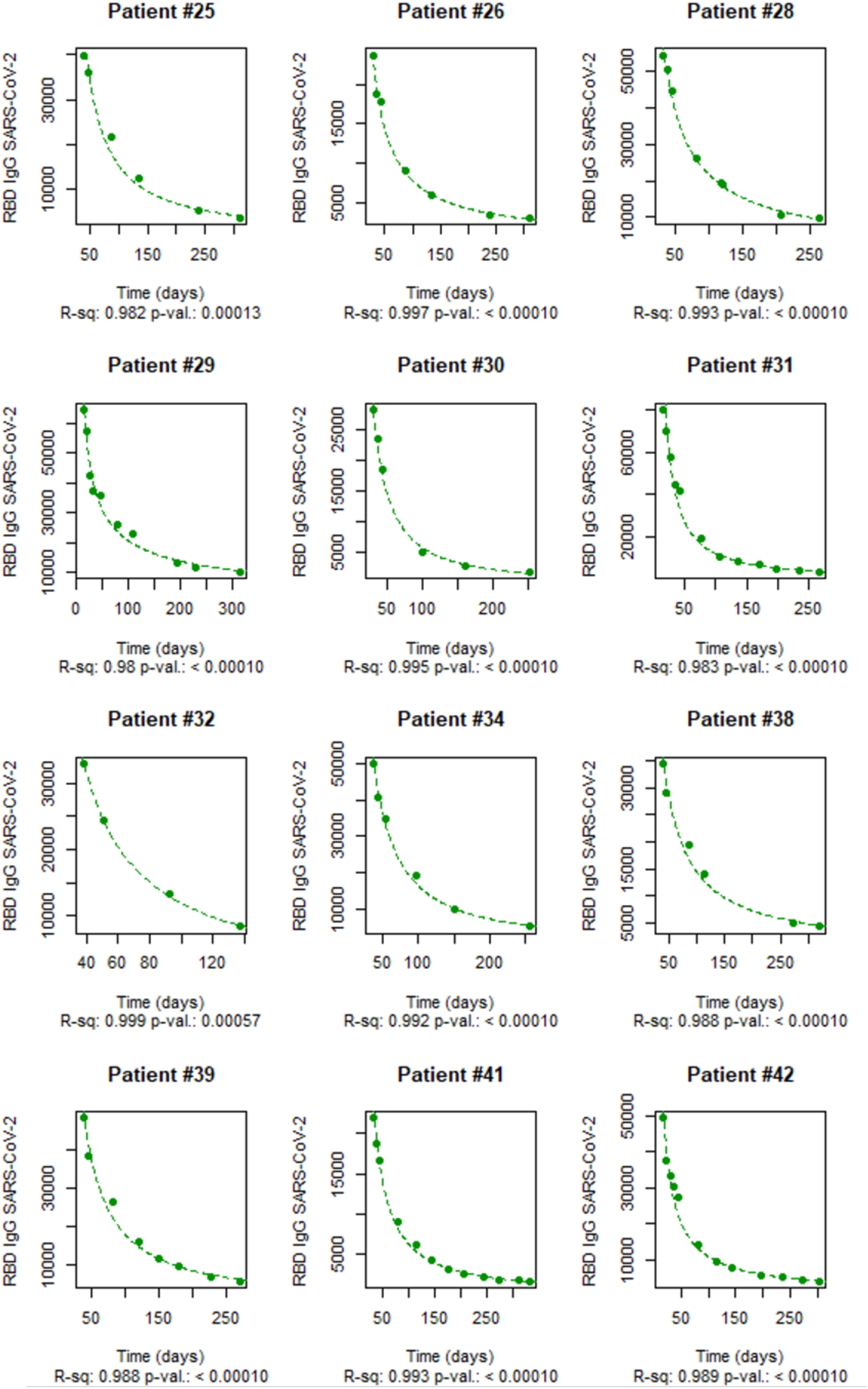
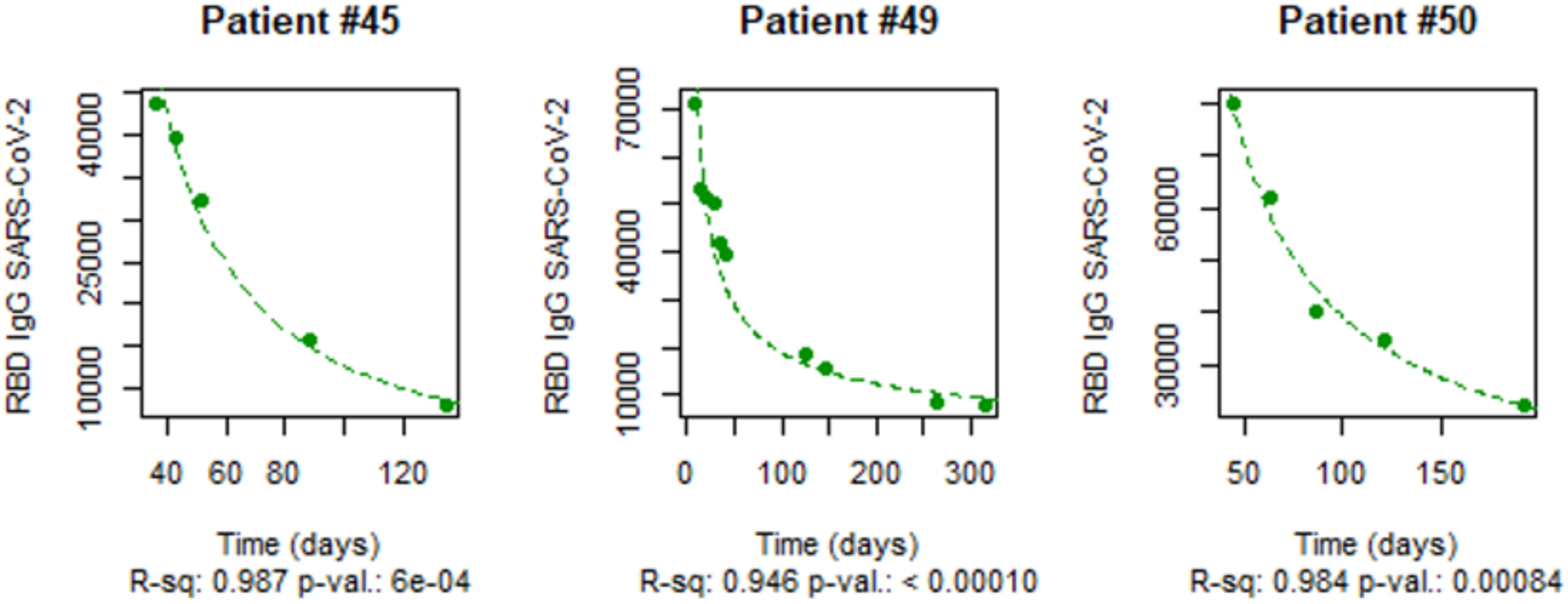
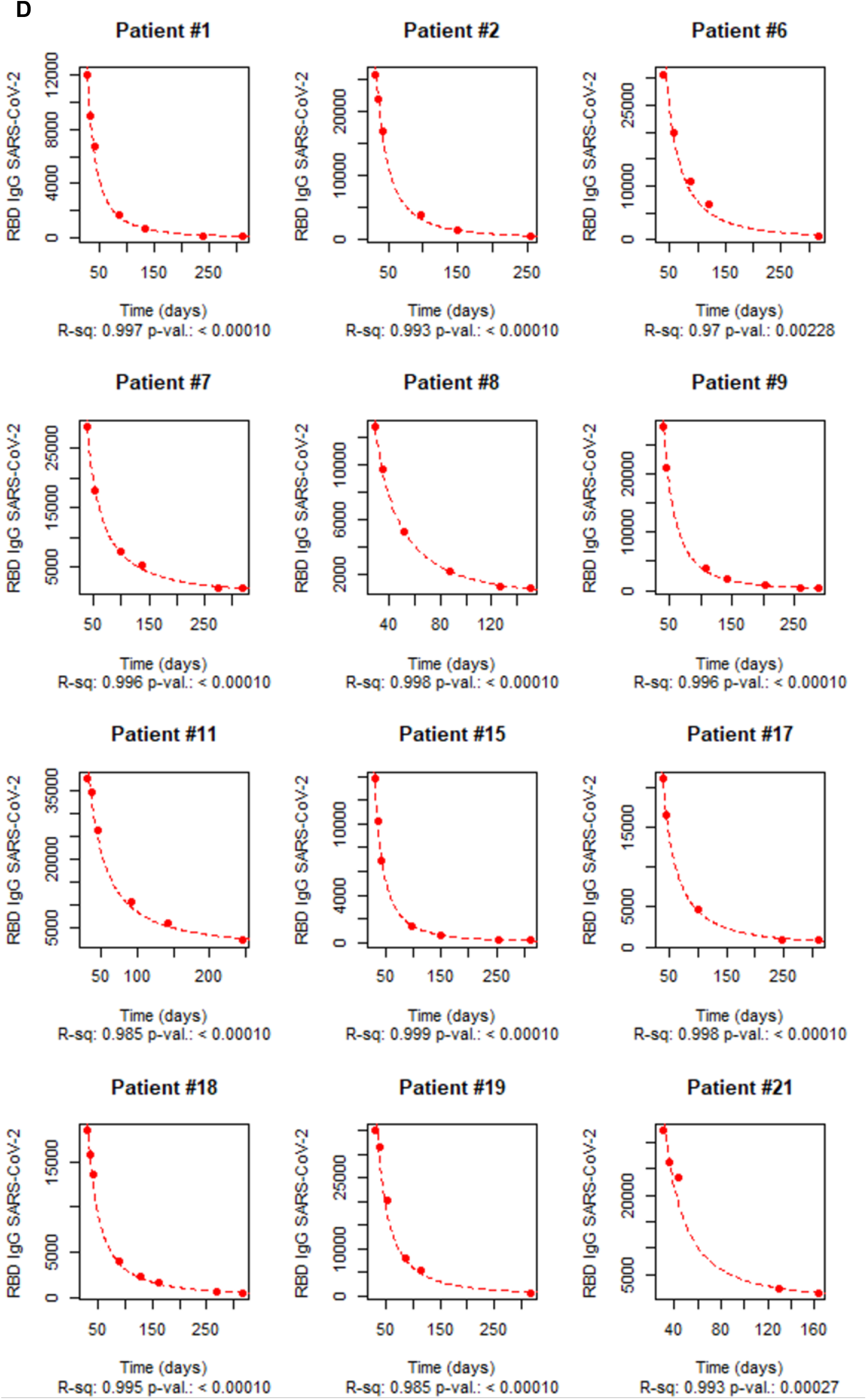
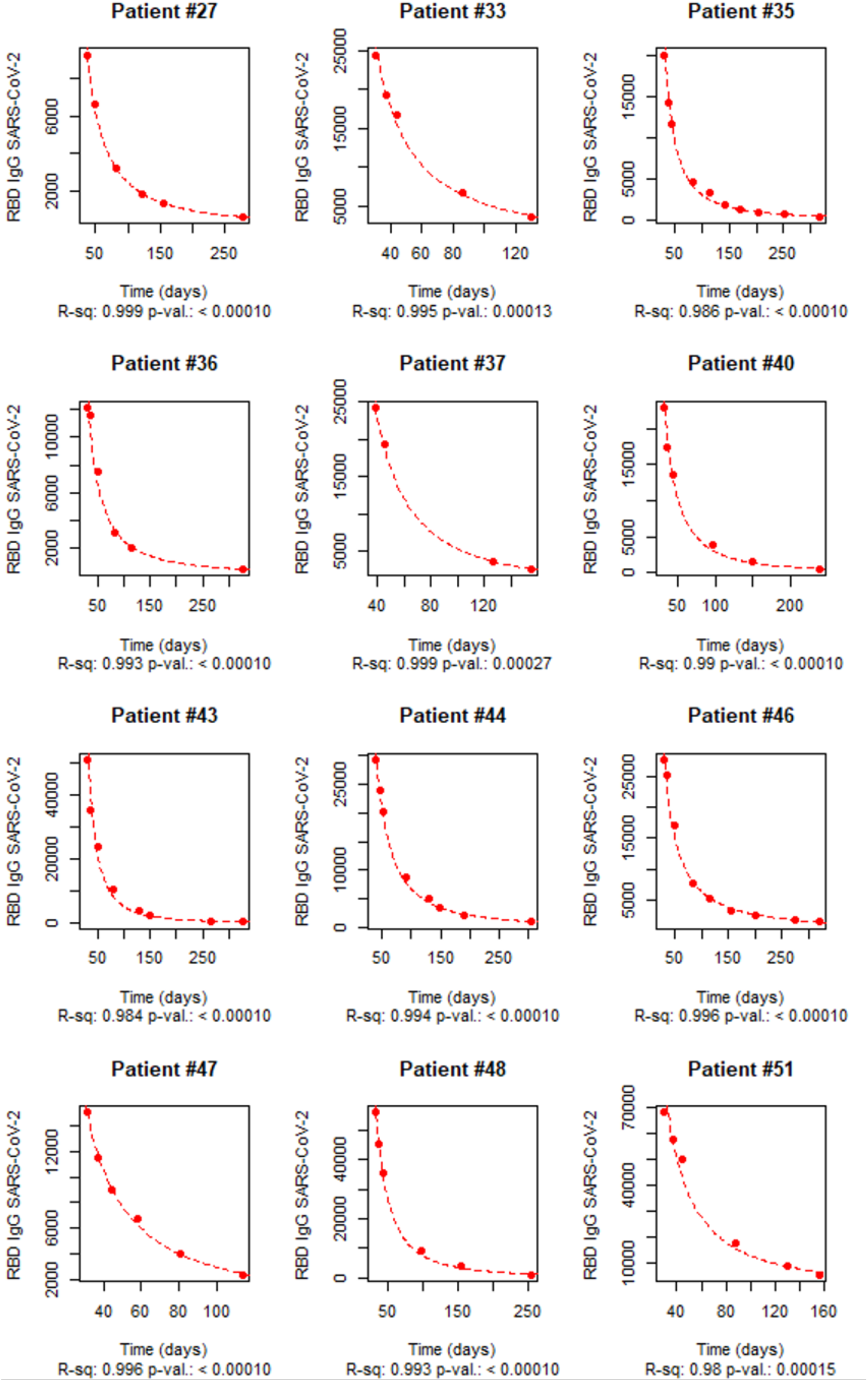
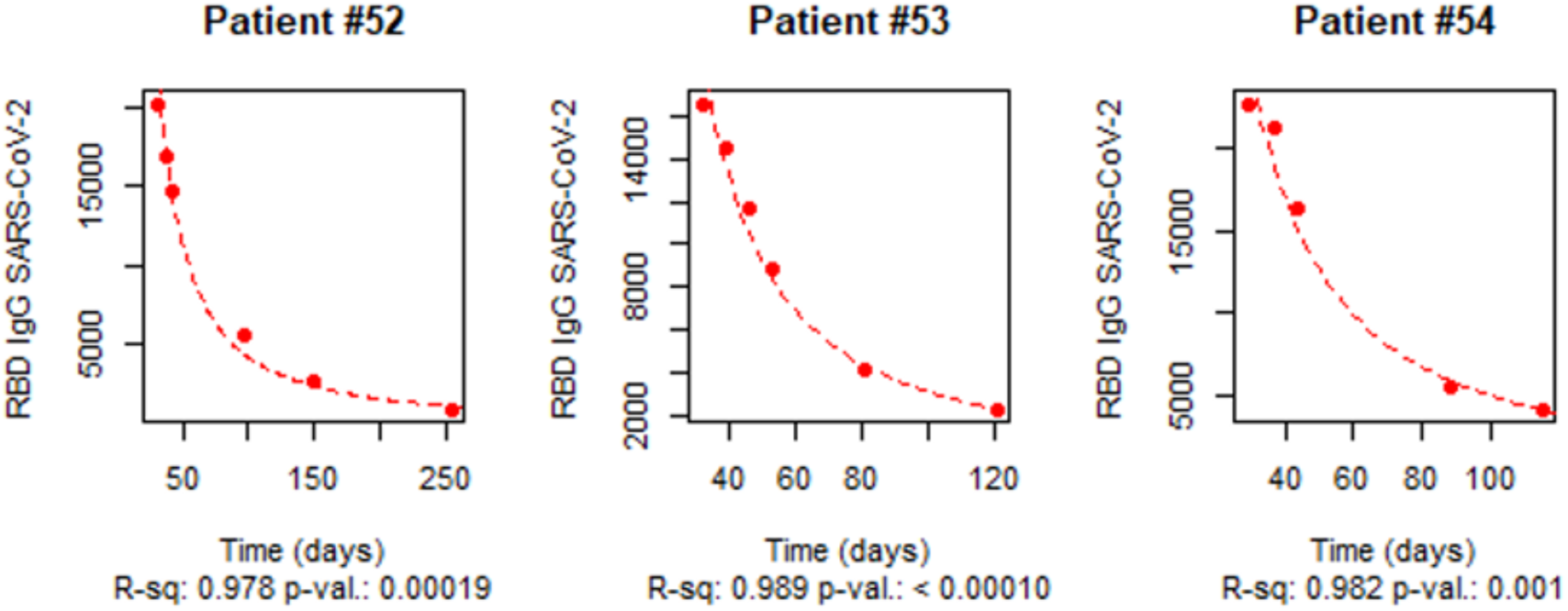
Time-course of anti-RBD IgG antibody titers for each individual HCW. A. Individual time-courses of naïve subjects adjusted by exponential model. B. Individual time-courses of experienced subjects adjusted by exponential model. C. Individual time-courses of naïve subjects adjusted by potential model. D. Individual time-courses of experienced subjects adjusted by potential model.

**Supp. 2.**
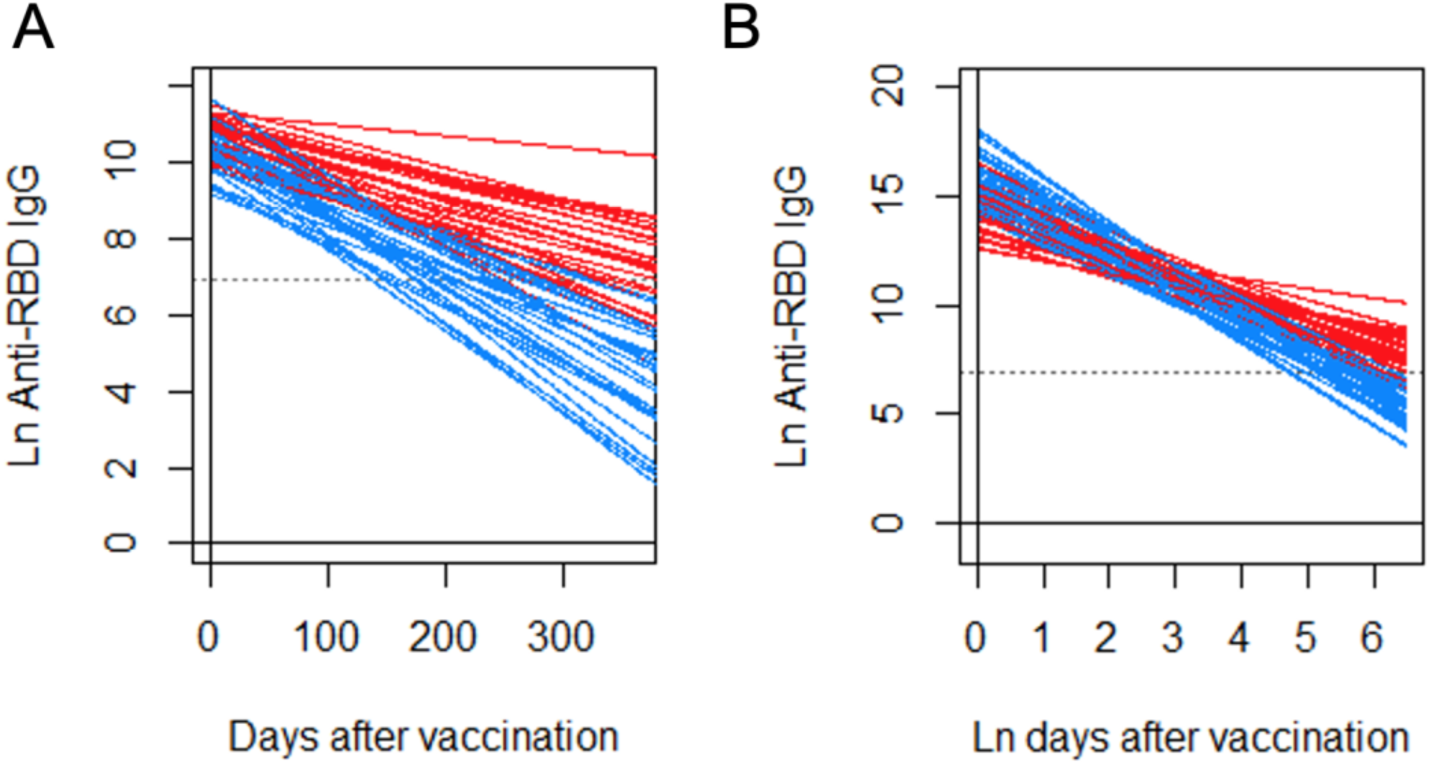
Linear transformation of individual curves for both nHCW (in blue) and eHCW (in red) groups for the exponential (A) and potential (B) models. The horizontal dashed line indicates 1.000 AU/ml.

**Supp. Table 1.**
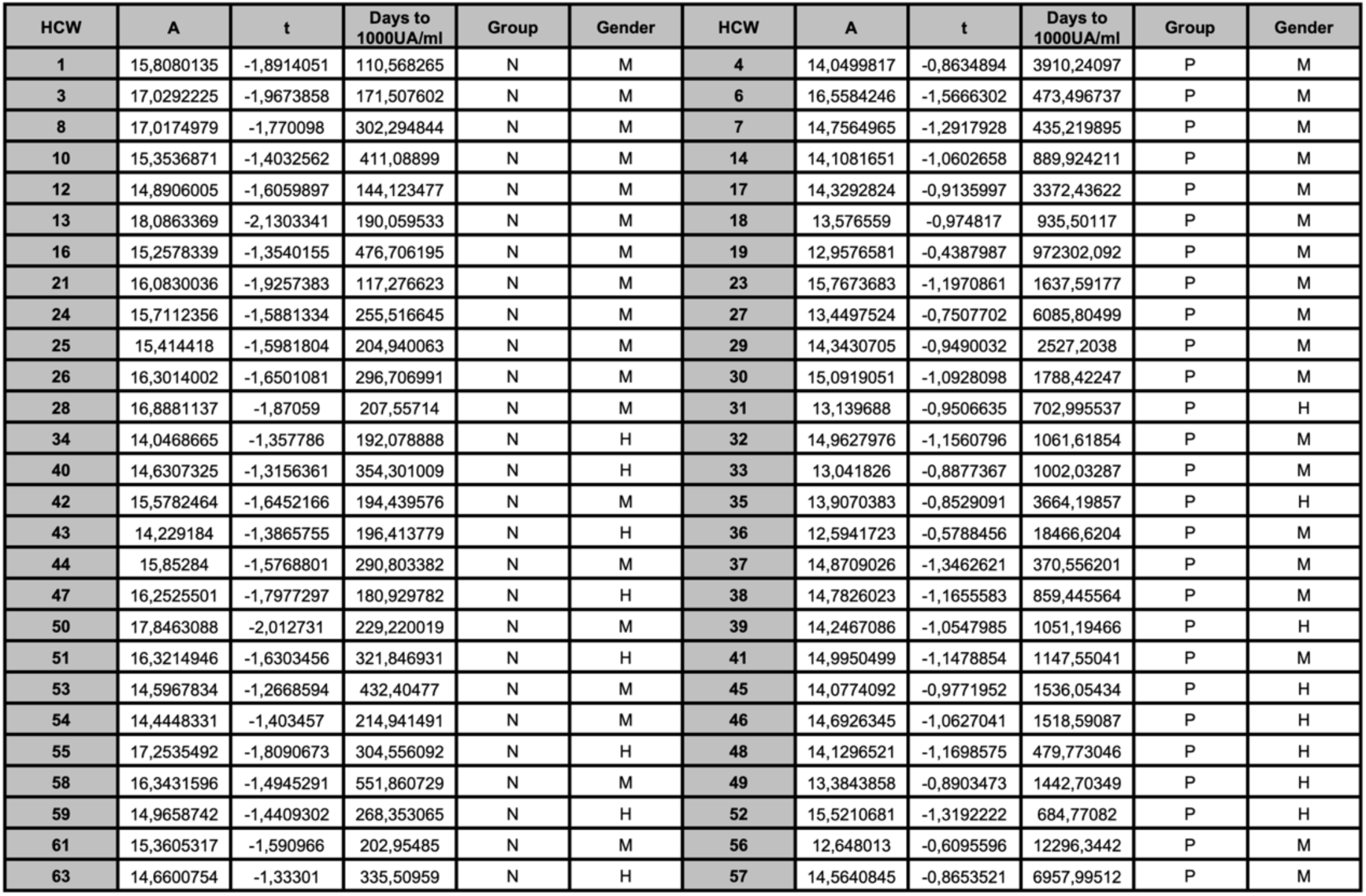
AIC comparison of exponential Vs. potential model.

**Supp. Table 2.** Statistics of the estimations of days in which each group would reach an anti-RBD IgG titer of 1000 UA/ml for the potential model in terms of mean, standard deviation (SD) and median. Due to the nature of the model, the estimation of the potential model for long periods of time develops an asymptotic behavior.

|  | nHCW (days) | eHCW (days) |
| --- | --- | --- |
| Mean | 265.1 | 38800.0 |
| SD | 108.0 | 186606.3 |
| Median | 229.2 | 1442.7 |

